# Assessing Knowledge About Pregnancy Induced Hypertension Among Pregnant Women Attending Antenatal Care at Makole Health Center

**DOI:** 10.1101/2024.07.24.24310928

**Authors:** Dennis Francis Mrosso, Baraka Dominick, Ally Machimu, Hassan Mwasi, Rehema Ramadhani, Tienyi Mnyoro Daniel

## Abstract

**Aim:** To assess the knowledge regarding pregnancy induced hypertension among pregnant mothers attending antenatal care at Makole health center.

**Methods:** Analytical cross-section study design involving 160 participants was conducted at Makole health center. Interviewer and self-administered structured questionnaire was used to assess the objectives of the study. coded, checked and then analyzed using Statistical Package for Social Science (SPSS) version 20 so as to develop descriptive statistical information presented inform of table, pie chart, histogram, and bar charts.

**Results:** Among 160 participants who were recruited in the study pregnant women aged 25 years or less constituted the majority 53(33.2%) with participants’ mean age of 22.61 with interquartile range of 18 to 43 years. Majority of the respondents had secondary education level 81 (50.6), while majority of the participants 111 (69.4%) were married. A greater proportion 101 (63.1%) of the pregnant women were Christians. With regard to occupational status, majority were self-employed 74(46.2%). While the majority of the respondents 126(78.8%) had low level of knowledge, 12(7.5%) had moderate level of knowledge and 22(13.8%) had high knowledge of pregnancy induced hypertension. The study found that the prevalence of PIH among pregnant women attending at Makole health center to be 8.1%.

**Conclusion:** Though the prevalence of pregnancy induced hypertension was low, a bigger proportion of these women did not have good knowledge of the disease and its complications. The study found that majority of those participants who were found to have experienced PIH had low level of knowledge. However, older pregnant women were ranging from moderate level of knowledge to high level of knowledge compared to the younger pregnant women whom majority of them had low level of knowledge. Also, health care providers should strengthen the awareness of pregnant women about pregnancy-induced hypertension in antenatal care clinics

## Background

Pregnancy Induced Hypertension (PIH) is a type of excessive blood pressure related to being pregnant. Pregnancy Induced Hypertension remains a number one purpose of high mortality and morbidity among ladies who are pregnant international (Ayele, Lemma & Agedew, 2016).Aside accelerated blood pressure ranges, PIH is likewise characterized by protein in urine (proteinuria) and peculiar edema (Tesfaye, Tefera. & Sena, 2018). The American College of Obstetricians and Gynecologists (ACOG) in collaboration with the United Nations (UN) Organization classifies hypertensive issues in pregnancy as: Chronic Hypertension, Gestational Hypertension, Preeclampsia/Eclampsia (Tebeu, Foumane, Mbu, Fosso, Biyaga & Fomulu, 2011). Pregnancy Induced Hypertension may be categorized as moderate or excessive. The former is while there’s a new onset of hypertension with the systolic blood pressure being ≥ a hundred and forty mmHg and or diastolic blood strain ≥ ninety mmHg, rising from 20 weeks’ gestation while the later diagnoses is made while blood stress measures ≥ 160mmHg systolic and ≥ a hundred and ten mmHg diastolic constantly (Ribowsky & Henderson 2012). Due to the severity of the circumstance in some women, high mortality is associated with PIH and is closely documented. Globally, approximately 3 hundred and fifty thousand (350000) women who are pregnant bypass away yearly from being pregnant associated reasons and over 50% of such Mortality taking area in Sub-Saharan Africa (SSA).It is expected that 12% of pregnant women’s deaths are related to PIH (Middendrop, Asbroek, Fred, Bio, Edusei, Meijjer, Newton & Agyemang, 2013).Globally, 10 % of pregnant girls are stricken by hypertension (Muti, Tshimanga, Notion, Bangure & Chonzi,(2015).A systematic overview with the aid of WHO concluded that PIH and its related complications is ranking 0.33 because the reason of maternal deaths in Africa, whereas in Latin America and the Caribbean, it contributes to 25.7% of mortality (WHO, 2011). In Africa, 9.1 % of mothers who die is due to hypertensive disorders of pregnancy (Arshad, Pasha, Khattak & Kiyani, 2011).

Aside the full-size mortality, PIH is linked with delivery earlier than time period, Intra Uterine Growth Retardation, Abruption Placentae and Intra Uterine Fetal Death (Muti, et al., 2015). In addition, complications springing up from PIH has the tendency of affecting each the mom and toddler (Jones, Takramah, Axame, Owusu, Parbey, Tarkang, Takase, Adjuik & Kweku, 2017). A range of chance factors have been documented to expect danger for PIH (Solomon & Seely, 2011). These encompass; Nulliparity, Multiple Pregnancies, History of Chronic Hypertension, Gestational Diabetes, Foetal Malformation and Obesity are associated with PIH (Khosravi, Dabiran, Lotfi & Asnavandy, 2014). Maternal age much less than 20 or over forty years, occurrence of PIH in preceding pregnancies, pre-current diseases Like kidney Disease, Diabetes Mellitus (DM), Cardiac sickness, undetected continual high blood pressure, fantastic family history of PIH in which there may be genetic vulnerability, mental, strain, use of alcohol, rheumatic arthritis, low Body Mass Index (BMI) and being overweight, and socioeconomic popularity being low is similarly link to PIH (Tesfaye, et al., 2018).

Maternal age much less than 20 or over forty years, incidence of PIH in preceding pregnancies, pre-present sicknesses Like Kidney Disease, Diabetes Mellitus (DM), Cardiac sickness, undetected persistent hypertension, nice own family records of PIH where there is genetic vulnerability, psychological strain, use of alcohol, rheumatic arthritis, low BMI and being obese, and socioeconomic fame being low are equally related to PIH (Tesfaye, et al., 2018).

Surprisingly, maximum girls aren’t knowledgeable on the life of PIH even as others have numerous perspectives on the physiological and pathological reason and others liaise its signs with superstitions (Brown, Best, Pearce, Waugh, Robson & Bell, 2013). In Sub-Saharan Africa, pregnant women were said taking the least suitable movements to reduce PIH as they typically maintain evil spirits and witchcraft answerable for the situation (Conde & Belizan, 2010). Evidence shows that alternative complementary medicinal drug changed into usually practiced with the aid of to preserve themselves from harm, alter the disease and to deal with high blood pressure (Conde & Belizan, 2010).

In a look at conducted inside the Tamale Metropolis, Ghana amongst ladies close to PIH, it become discovered that 60% of the observe contributors lack information on self-care control of PIH (Muzakiel, 2013). In addition, a similar take a look at conducted by way of Musah and Iddrisu (2013) inside the Tamale Teaching Hospital found out that about forty% of pregnant girls who attended ANC lack expertise of self-care control of PIH and some of the pregnant women even considered edema and weight advantage as everyday situations of pregnancy. Hence, understanding approximately PIH and its control is important as it impacts fitness looking for behavior among pregnant women in the world (Tuovinen, Raikkonen & Pesonen, 2012).

In Tanzania, a study conducted in a District Hospital in Tanzania indicated that 60% of the study individuals did not recognize the outcomes of preeclampsia. Lack of expertise is observed to be the predisposing issue to practice risky behaviors for preeclampsia. Moving greater closer the superiority of excessive PIH amongst postpartum girls in Zanzibar is high about 26.3%. Common threat elements on this placing encompass loss of popular information on PIH, maternal age of 15–20years, own family history of high blood pressure, pre-current diabetes previous to idea, and multifetal gestation (Machano & Joho BMC Public Health, 2020).Another observe became carried in Makole Dodoma concerning knowledge on PIH in which 200 adult girls had been recruited in the diagnosed community, the general expertise ranges were low with a mean of 41% of accurate solutions (Savage & Hoho, 2016).

## Methods

### Study design and Setting

This is an analytical cross-sectional study carried out at Makole health center. Makole health center is a public facility that provides surgical, medical and basic diagnostic healthcare services of high quality, effective and accessible to all, delivered by well performing and sustainable national health system, it located in Dodoma region. The region is located in central zone of Tanzania and covers 41,311 km^2^ with a total population of around 2,083,588 people. On east side it is nearby Morogoro region, northward is nearby Manyara region, on westward is nearby Singida region and on southward is nearby Iringa region. Also, the city is divided into seven districts; Dodoma urban, Bahi, Chamwino, Kondoa, Mpwapwa, Kongwa and chemba. The districts are sub divided into 209 wards and 637 registered villages. The economy of Dodoma is based on subsistence agriculture and animal husbandry (National Bureau of Statistics, 2013).

### Study population

All pregnant women attending ANC at Makole Health Center with a gestational age of 20 weeks and above who were accessible during the study period and gave their consent to be part of the study were included as study participants, while all referrals at the time of data collection and pregnant women who were unable to communicate or who were critically ill, as well as those who declined to participate in the study, were excluded from the study.

### Sampling and Recruitment procedure

Simple random sampling method was used to select participants where a sampling frame from all pregnant women with 20 weeks’ gestation or above was developed from daily ANC attendance.

### Sample size determination

The sample size for the study was computed by using Cochran’s formula. Based on this, the maximum sample size was calculated as follows:

From the Cochran’s formula;

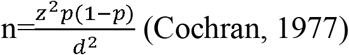

Where:

n= required sample size

Z^2^ = standard normal deviate for two tailed tests based on 95% confidence level = 1.96

p= proportion of pregnant women with pregnancy induced hypertension = 28.5% = 0.285

q=1-p= proportion of pregnant women without pregnancy induced hypertension= 1-0.285= 0.715 e= margin of error = 5% = 0.05

Therefore, the sample size will be calculated as follows

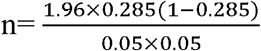

n= 159.7596 = 160 participants

Therefore, 160 pregnant women required in this study.

### Data collection method

Data was collected through both interviewer and self-administered structured questionnaire with open and close ended questions to collect information from study participants who attended ANC at Makole health center. The open-ended questions used to allow respondents to provide their response with more details. And close ended questions enabled respondents to select the best option among those prepared for each question. The questionnaire designed with questions that will intend to explore about demographic data and knowledge of pregnancy induced hypertension. The questionnaire was prepared in English then translated into Swahili for easy understanding, then it was translated back into English to assess for consistency. The tool was tested on 10% on the sample size in a town that was selected for actual study and modified accordingly. The reliability of the questionnaire was calculated based on data from the pretest.

### Data analysis

Data collected was coded, checked and then analyzed using Statistical Package for Social Science (SPSS) version 20 so as to develop descriptive statistical information presented inform of table, pie chart, histogram, and bar charts.

### Ethical consideration

Study was detailed examined and approved ethically by Internal Ethics Review Committee(IERC) of St. John’s University of Tanzania on 15^th^June, 2022. Permission to conduct this study was authorized by the district medical officer (DMO). Approval was also sought from the medical officer and head of the antenatal care clinic at the Makole health Center. Participants were thoroughly informed about the objective of the study and assured that participation is purely voluntary and they can opt out at any time, and this would not affect service delivery at the facility. The data collected was kept under lock and key, with only the principal investigator having access. To ensure anonymity, participants were only identified with numbers instead of their actual names during and after data collection.

## Results

### Socio-Demographic Characteristic of the Respondents

A total 160 pregnant women were recruited as participants of the study. Pregnant women aged 25 years or less constituted the majority 53(33.2%) with participants’ mean age of 22.61 with interquartile range of 18 to 43 years as shown in table 1. Majority of the respondents had secondary education level 81 (50.6), while majority of the participants 111 (69.4%) were married. A greater proportion 101 (63.1%) of the pregnant women were Christians. With regard to occupational status, majority were self-employed 74(46.2%) as shown on table 1.

**Table 1:** social demographic characteristics of respondents (N=160)

| VARIABLE | FREQUENCIES | PERCENTAGE (%) |
| --- | --- | --- |
| **Age** |  |  |
| <25 | 53 | 33.2 |
| 25-29 | 43 | 26.9 |
| 30-34 | 42 | 26.2 |
| 35-39 | 20 | 12.4 |
| 40-45 | 2 | 1.2 |
| **Marital status** |  |  |
| Married | 111 | 69.4 |
| Single | 36 | 22.5 |
| Divorced | 1 | 0.6 |
| Single parent | 12 | 7.5 |
| **Education level** |  |  |
| No formal education | 6 | 3.8 |
| Primary education | 34 | 21.2 |
| Secondary education | 81 | 50.6 |
| College/University | 39 | 24.4 |
| **Religion** |  |  |
| Christian | 101 | 63.1 |
| Muslim | 46 | 28.8 |
| Others | 13 | 8.1 |

### Knowledge on Pregnancy Induced Hypertension and its related complications

Majority of the participants had low level of knowledge 126(78.75%) regarding pregnancy induced hypertension as shown on figure 2. The most common identified signs and symptoms of the disease condition were swelling of the face, hands and legs 35(21.9%), high blood pressure 26(16%), dizziness 35(21.9%), headache 30(18.8%), abdominal pain 11(6.9%), itching 18(11.2%), vomiting and diarrhea 7(4.4%) and blurred vision 27(16.9%) as shown in Table 2.

**Figure 1.**
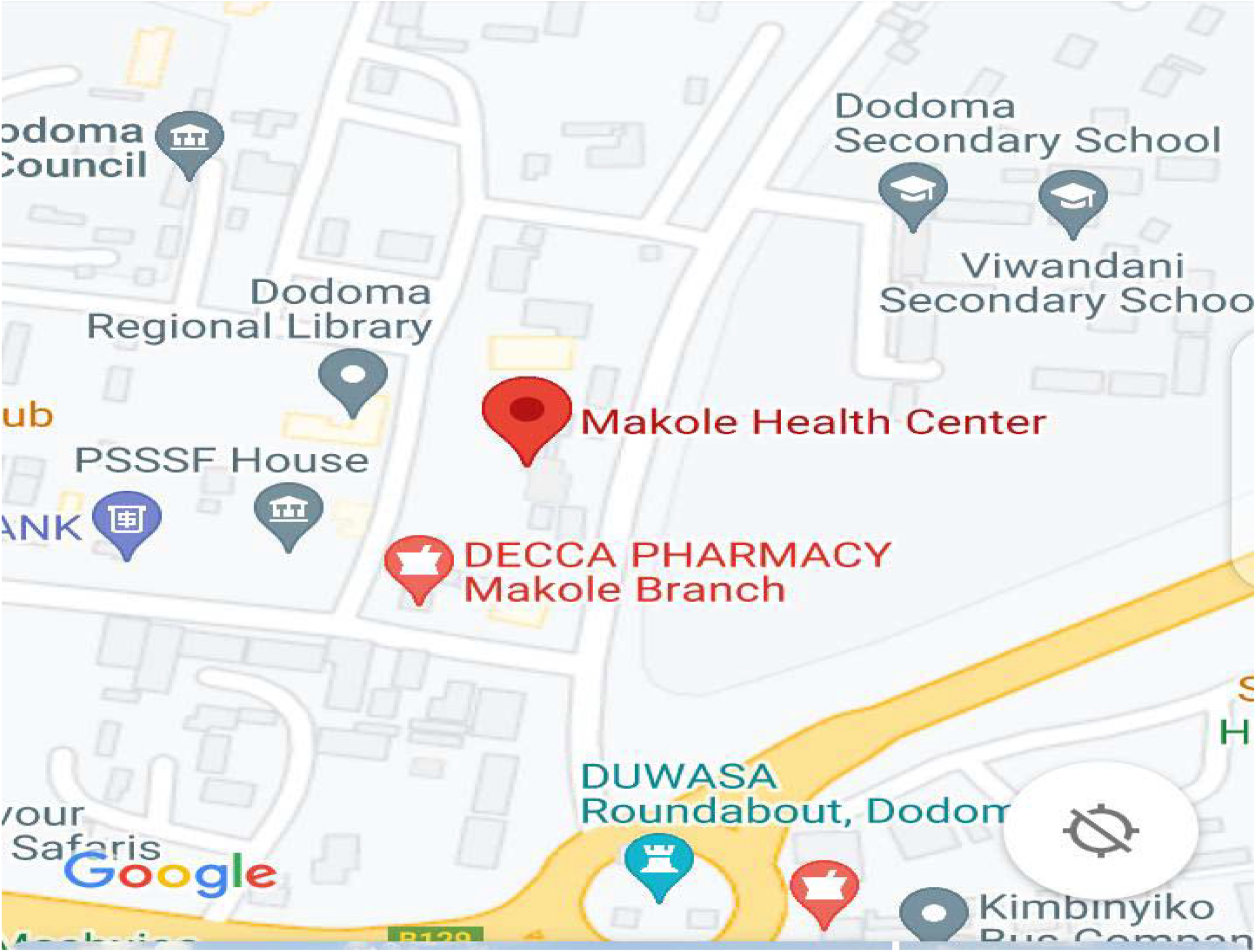
location of Makole health Center. **Source:** Google maps: https://www.google.com/maps/place/Makole+health+center/

**Figure 2:**
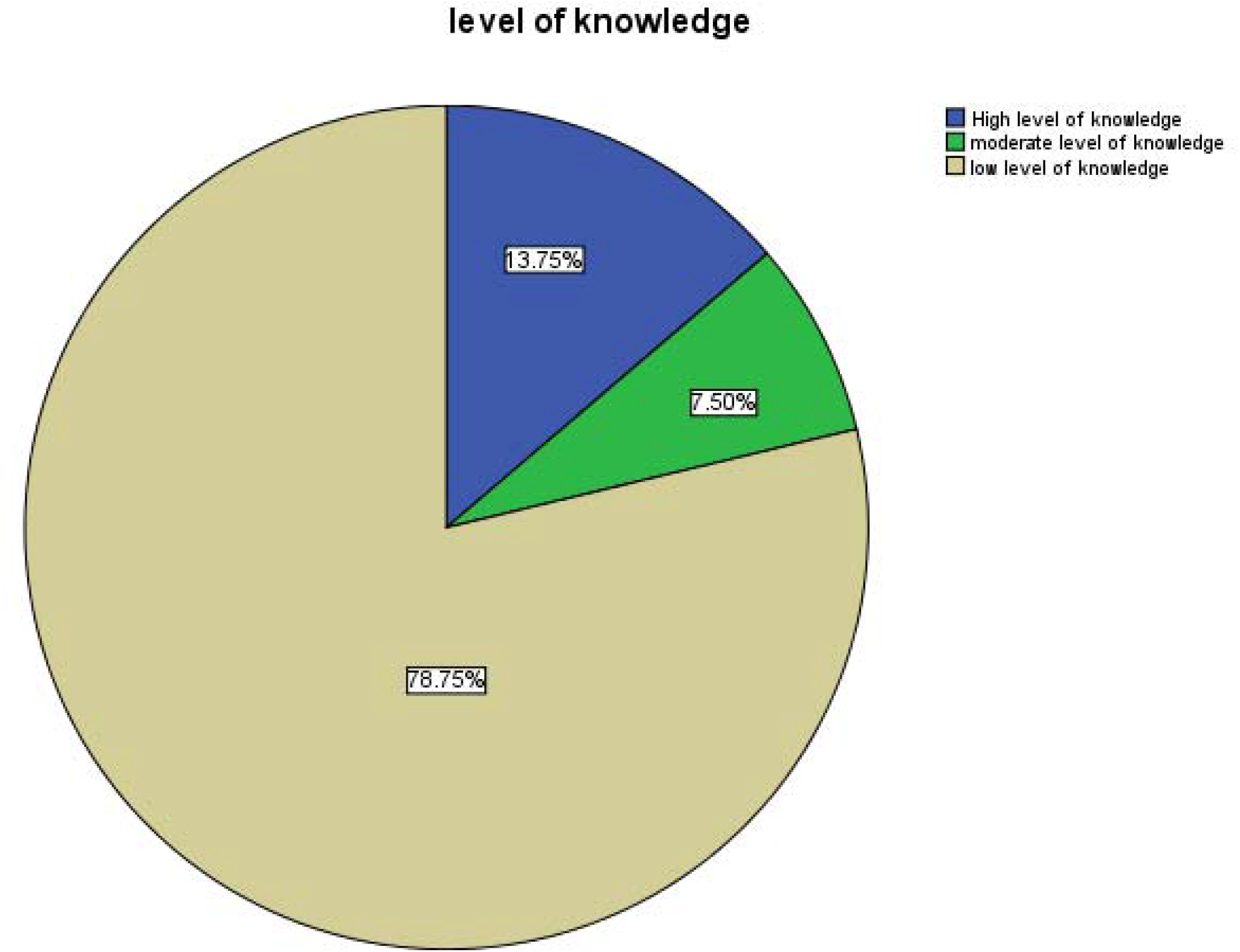
Knowledge on pregnancy induced hypertension.

**Table 2:** Knowledge on Pregnancy Induced Hypertension and its related complications.

| VARIABLE | RESPONSE | FREQUENCIES | PERCENTAGE (%) |
| --- | --- | --- | --- |
| **Statements** |  |  |  |
| Swelling of the face, hands and legs | *NO* | 1 | 6 |
|  | *YES* | 35 | 21.9 |
| Abdominal pain | *NO* | 10 | 6.2 |
|  | *YES* | 11 | 6.9 |
| Dizziness | *NO* | 1 | 6 |
|  | *YES* | 35 | 21.9 |
| Vomiting and diarrhea | *NO* | 20 | 12.5 |
|  | *YES* | 7 | 4.4 |
| Itching | *NO* | 10 | 6.2 |
|  | *YES* | 18 | 11.2 |
| Headache | *NO* | 4 | 2.5 |
|  | *YES* | 30 | 18.8 |
| High blood pressure | *NO* | 6 | 3.8 |
|  | *YES* | 26 | 16 |
| Blurred | *NO* | 4 | 2.5 |
|  | *YES* | 27 | 16.9 |

### Prevalence of PIH

The study found prevalence of PIH among pregnant women attending at Makole health center to be 8.1%. as summarized in table 3.

**Table 3.** Prevalence of pregnancy induced hypertension among participants.

| **Variable** | **Response** | **Frequencies** | **percentage(%)** |
| --- | --- | --- | --- |
| Ever experienced have hypertension during pregnancy | YES | 13 | 8.1 |
|  | NO | 147 | 91.9 |

## Discussion

### Socio-demographic characteristics of the respondents

From the study, 160 participants who were recruited had a mean age of 22.61 with interquartile range of 18 to 43 years, this is encouraging as this is active reproductive age group and most of the respondents were married 111(69.4%). These background information are similar to those reported in a study conducted in Tigray, Ethiopia (Berhe et al., 2020). With respect to education, religion and occupation findings; majority of the respondents had secondary education level 81 (50.6%) while greater proportion 101 (63.1%) of the pregnant women were Christians. With regard to occupational status, majority were self-employed 74(46.2%). A similar study conducted in Mizan-University teaching hospital and other areas reported majority of respondents 150(36.1%) had primary education, 236(56.7%) were Christian and 276(66.3%) were housewives respectively (Tesfaye et al., n.d.)

### Prevalence of PIH

From the study, among 160 recruited participants the prevalence of PIH among pregnant women attending at Makole health center was 8.1%. This observation is close to those reported in a study on prevalence and determinants of pregnancy induced hypertension among women attending antenatal clinic at 37 military hospitals in Accra which was 8.8% (Marbell C.C, 2019). Similarly, a cross sectional study done in parts of Ethiopia reported prevalence of PIH of 7.9% (Tesfaya & Tilahun, 2019). Likewise, a systematic review study done in Ethiopia found prevalence of PIH was 6.07% (Berhe et al., 2018). These figures are not widely interspaced indicating nearly similar prevalence of PIH among pregnant women. These concurrences may be attributed with the fact that most Sub-Saharan African countries share common life styles. However, a cross-sectional study conducted in Korle Bu, Ghana recorded a prevalence of 21.4% (Adu-Bonsaffoh, 2017). This prevalence appears to be extremely higher than the prevalence from this and other studies (Marbell C.C, 2019, Tesfaya & Tilahun, 2019; Berhe et al., 2018). The variation in prevalence of PIH could be associated with variation in biological factors such as having family history of PIH, gestational age and the presence of ailments such as asthma and kidney problems among pregnant women as suggested by (Tesfaya & Tilahun, 2018). The difference in prevalence of PIH could also be attributed to differences in sample size and population size of the other study settings. Regardless, there is still a significant burden of PIH in the Tanzania population as evidenced by these prevalence rates hence, the need for more action-oriented research to determine cause and solutions to this problem.

### Knowledge of pregnancy induced hypertension among pregnant women and its related complications

Knowledge level on PIH Among the participants was assessed with the following findings; 13.75% had high level, 7.5% moderate and 78.75% low level of knowledge on the subject matter. These findings are less or more similar to those reported in a study conducted in India among primigravida mothers in Saveetha Medical college and hospital which reported 40% had average good knowledge, 15% had good knowledge and 5% had poor knowledge respectively (Muthulakshmi & Sowmiya, 2019). The current findings contrast those reported in a study conducted in Nigeria among health care workers in a Maternity Hospital which revealed the following on knowledge outcomes ; 14.5% had highest level and 16.4% had low level (Olaoye et al., 2019). This variation may be attributed to the fact that the later study was done on health professionals (Nurses, physicians’ etc.) who had good theoretical background on PIH but just lacked management experiences as observed. Furthermore these findings are comparable to earlier study conducted in the same setting with regard to the subject matter (Savage & Hoho, 2016). This observation implies that health education in regard to PIH is still needed to most pregnant women on prevention and management of this health problem.

In addition, participants were able to identify at least some signs and symptoms of PIH such that 36(22.5%) of pregnant women reported that knew the signs and symptoms of pregnancy induced hypertension. The most common identified signs and symptoms of the disease condition being swelling of the face, hands and legs 35(21.9%), high blood pressure, dizziness, headache, abdominal pain, itching, vomiting and diarrhea and blurred vision. These findings are similar with those identified in as study conducted in Ethiopia (Berhe et al., 2020), where headache 92 (29.4%),loss of consciousness 33 (10.5%) and new onset of visual disturbances. Similar findings have been reported from studies as stipulated in a systematic review of Sub-Saharan Africa (Wagnew et al., 2020). Likewise, concurrent observation have been reported in a study conducted in Sri-Lanka on maternal and fetal effects at tertiary care hospital (Dodampahala et al., 2014). These similarities may be attributed to the fact that these studies were all done in African settings where the participants share most of the life style as well as culture.

## CONCLUSION

The study found the prevalence of PIH to be 8.1% among pregnant women seeking care from Makole health center. Results of this and other studies have suggested that women who encounter Pregnancy Induced Hypertension (PIH) are greatly challenged with pregnancy outcomes including maternal mortality. Broadly, majority of the pregnant women are ill-informed about PIH including its signs and symptoms, complications and management. Apparently, an improvement in knowledge on PIH among pregnant women will lead to an improvement in early reporting and management of PIH cases in early stages. The role of health care providers in health education and early management of PIH among pregnant women cannot be underestimated as these forms the baseline for reducing complications arising from PIH. It therefore implies that there should be increased human resources, capacity building and in-service training of staff on proper management of PIH.

## RECOMENDATION

- The ministry of health and other stakeholders should put much emphasis on health promotion initiatives and mass education to women of reproductive age to ensure that most community members become well aware of all aspects regarding pregnancy induced hypertension as means of primary disease prevention involving early screening and other health promotion activities as it found to be more effective in increasing awareness, decrease morbidity and mortality related to hypertension in pregnancy.
- Staff at the Antenatal care (ANC) clinics should periodically organize health education topic on PIH including its causes, signs and symptoms, complications and the need for immediate reporting to the facility anytime they (pregnant women) observe any PIH signs and symptoms and use posters in local language as well so as to increase awareness. This will breach the gap in knowledge and an improvement in reporting of the condition by pregnant women.
- Pregnant women should be emphasized to attend these health educational sessions of ANC for their own benefits. Family members, especially husbands, should also participate in these sessions in order to encourage their pregnant women to comply with the issues discussed.

## Data Availability

All data produced in the present study are available upon reasonable request to the authors

## Declarations

### Consent for publication

Not applicable.

## Author’s contributions

**D.M** developed a study protocol, conducted data collection, performed data analysis and drafted a manuscript.

**B.D** developed a study protocol, conducted data collection, performed data analysis and drafted a manuscript.

**A.M** developed a study protocol, conducted data collection, performed data analysis and drafted a manuscript.

**H.M** developed a study protocol, conducted data collection, performed data analysis and drafted a manuscript.

**R.R** developed a study protocol, conducted data collection, performed data analysis and drafted a manuscript.

**T.D** Reviewed and develop study protocol, survey tool and drafted manuscript.

## Availability of data

Not applicable

## Acknowledgment

The authors would like to appreciate support from the St. John’s University of Tanzania for full support in accomplishment of this study.

